# A Randomized Hybrid Type I Effectiveness-Implementation Study of an eHealth Delivery Alternative for Cancer Genetic Testing for Hereditary Cancer (eREACH2): study protocol

**DOI:** 10.1101/2025.11.19.25340515

**Authors:** E Mastaglio, B Egleston, KT Lee, D Fetzer, S Brown, SM Domchek, Linda Fleisher, KY Wen, L Wagner, JS Roberts, C Cacioppo, J Christiansen, S Howe, EM Wood, M Weinberg, K Karpink, E Selmani, J Feng, S John, K Schweickert, B McLeod, AR Bradbury

## Abstract

**Background:** Germline cancer genetic testing has become a standard evidence-based practice, with established risk reduction and cancer screening guidelines for genetic carriers. There are also targeted treatments for some genetic carriers with cancers such as breast and ovarian cancer. Yet, many at-risk patients do not have access to genetic services, leaving many genetic carriers unidentified. The eREACH 2 study (A Randomized Hybrid Type I Effectiveness-Implementation Study of an eReach Delivery Alternative for Cancer Genetic Testing for Hereditary Cancer) evaluates whether an interactive, patient-centered digital alternative for genetic education and disclosure of results is non-inferior compared to the traditional model of pre-and post-test counseling with a genetic counselor.

**Methods:** This is a Hybrid Type 1 effectiveness implementation study in which participants are randomized using a 2×2 design to test a self-directed, patient-informed, digital intervention to deliver clinical genetic testing versus the traditional pre-test (visit 1) and post-test (visit 2: disclosure) counseling delivered by a genetic counselor. The 4 arms include A) genetic counselor for visit 1 and visit 2, B) genetic counselor for visit 1 and digital intervention for visit 2, C) digital intervention for visit 1 and genetic counselor for visit 2, and D) digital intervention for both visits. Participants are adults who meet National Comprehensive Cancer Network and/or American Society of Clinical Oncology guidelines for germline genetic testing and are recruited from both community and medical sites across the United States by way of clinician referral as well as patient self-referral. The primary outcomes are non-inferiority in uptake of genetic testing and change in genetic knowledge and general anxiety from baseline to post-disclosure.

**Discussion:** With many barriers to accessing genetic services, innovative delivery models are needed to address these gaps and increase uptake of genetic services. The eREACH2 study evaluates the effectiveness of an interactive patient-centered digital intervention to deliver clinical genetic testing. We expect this work will inform evidence-based guidelines and the standard-of-care for delivery of genetic testing and is designed to be broadly applicable and easily adaptable for other populations and settings even beyond oncology (e.g. Alzheimer’s disease).

**Trial registration:** This protocol was registered at clinicaltrials.gov (NCT05427240) on 6/7/2022.

## BACKGROUND

There are established risk reduction and cancer screening guidelines for cancer genetic carriers.(1–4) Germline cancer genetic testing also has the potential to personalize cancer treatment with the approval of treatments such as poly adenosine diphosphate ribose polymerase (PARP) inhibitors for patients with germline *BRCA1* and *BRCA2* mutations and breast, ovarian, pancreatic, and prostate cancers.(5, 6) Yet, many at-risk patients do not have access to genetic services, leaving many genetic carriers unidentified.(7–11) The current workforce of genetic specialists is not sufficient to meet current and risking indications for genetic testing under the traditional model of pre-and post-test counseling with a genetic professional.(12, 13) Thus, there is an urgent need to consider alternative delivery models to increase access and uptake of genetic testing, while maintaining positive patient outcomes such as knowledge and psychological adaptation to test results.

Several studies have identified favorable outcomes with remote provision of genetic counseling services either by phone of videoconference.(14–21) While remote telegenetic services can increase access to services, this model alone does not address the growing indications for clinical genetic testing and the limited genetic counselor (GC) workforce. Biesecker and colleagues reported that a web-based platform was non-inferior to provider-mediated genetic counseling in a highly educated cohort undergoing carrier testing, and the RESPECT study demonstrated that a web-based pre-disclosure education intervention in lieu of standard pre-disclosure counseling with a GC was acceptable to participants with 88.5% of participants opting for web disclosure.(22, 23) The recent eREACH study found that digital delivery of pre-test counseling or post-test disclosure of results was non-inferior to two visits with a GC among patients with certain metastatic cancers.(24) Thus, interactive, patient-centered digital interventions, used in collaboration with genetic specialists may be an alternative way of addressing the workforce shortage while maintaining adequate patient outcomes.(14, 23–25)

### Conceptual Model

Our studies evaluating delivery innovations in genetic services have been informed by our conceptual model grounded in The Self-Regulation Theory of Health Behavior (SRTHB, **Figure 1**).(25–28) It proposes that the *<u>reaction to</u>* and *<u>use of</u>* health (genetic) information is the product of an individual’s *<u>understanding</u>*, (i.e. knowledge and perceptions of risk) and the biopsychosocial impact of the risk and the health behavior.(26, 29–33) It emphasizes “common-sense” representations rather than medical or scientific definitions, and incorporates individual biological, cognitive, emotional, familial and cultural experiences that might impact variability in understanding and risk modification for individuals and their families. Thus, it has been proposed that the SRTHB is an ideal framework for considering the outcomes and effectiveness of genetic testing, particularly among diverse patient populations.(27, 31, 34)

**Figure 1:**
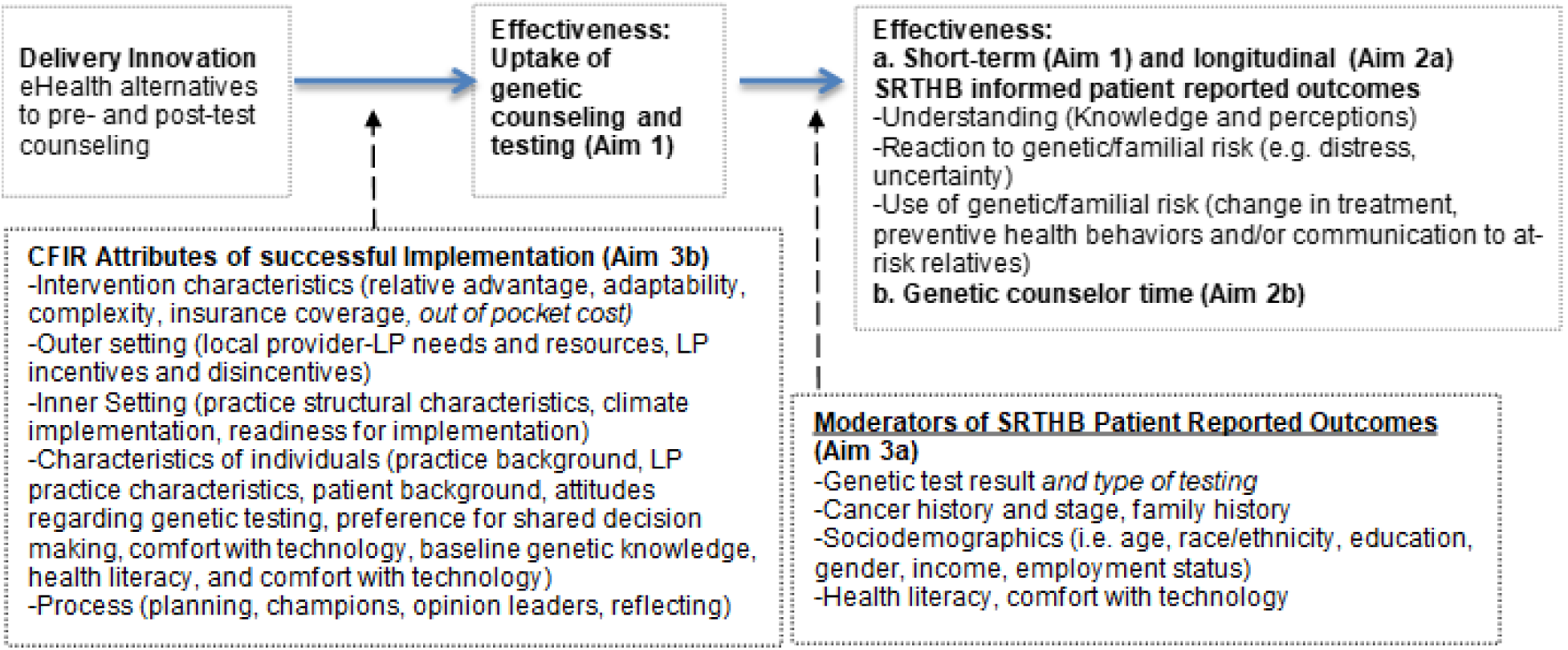
CONCEPTUAL MODEL to evaluate innovations to delivery of genetic services: guided by the Self-Regulation Theory of Health Behavior (SRTHB) and Consoidated Framework for Implementation Research (CFIR)

Even after health-related interventions have proven efficacy and effectiveness, many fail to translate into clinical settings.(35, 36) Thus, there is increasing recognition of the importance of evaluating and addressing barriers to implementation across diverse health care settings. The Consolidated Framework for Implementation Research (CFIR) provides an overarching theoretical framework to evaluate barriers, enhancements and adaptations to increase successful implementation of effective interventions and delivery adaptations.(37) We have included constructs from each of the 5 major CFIR domains (intervention characteristics, outer setting, inner setting, characteristics of individuals and process) as they relate to our intervention (see **Table 1)**, which will be evaluated in our multi-stakeholder mixed-methods process evaluation. As in other studies, evaluation of CFIR components is expected to identify patient, provider and system barriers and enablers of our eHealth delivery alternatives and increase future dissemination to increase access to, and uptake of genetic services, while maintaining optimal patient outcomes. (38–40)

**Table 1:** Implementation Outcomes and Measures based on the Consolidated Framework for Implementation Research.

| Measures | Sample Items | Local provider registration | Patient Survey | Telegenetics service records | Key Informant Interviews |
| --- | --- | --- | --- | --- | --- |
| Intervention Characteristics |  |  |  |  |  |
| Relative advantage | Perception that telehealth and digital alternative service options provides an advantage over existing care options | X | X |  | X |
| Adaptability | Perception that telehealth and digital alternative service options can be modified to meet practice, provider or patient needs, recommendations for improvement | X | X |  | X |
| Complexity | Perception that telehealth and digital alternative service options are too complex | X | X | X | X |
| Cost | Perception that telehealth and eHealth alternative service options or genetic testing would be too costly |  |  | X | X |
| Outer setting |  |  |  |  |  |
| Local provider, genetic counselor and insurer needs and resources | Awareness of genetic risk and cancer prevention; importance to practice or genetic counseling field | X |  |  | X |
| Provider incentives and disincentives | External mandates for genetic testing (e.g. accreditation); Genetic counselor benefits, disincentives and ways to address benefits and disincentives to using digital alternatives | X |  |  | X |
| Inner Setting |  |  |  |  |  |
| Structural characteristics | EMR, support staff characteristics and needs, practice structure | X |  | X | X |
| Climate for implementation | Previous innovation implementation, compatibility with practice, relative priority, quality metrics/rewards | X |  | X | X |
| Readiness for Implementation | Available staff resources, practice experience with telehealth and digital genetic services | X |  | X | X |
| Characteristics of Individuals |  |  |  |  |  |
| Practice background | Type of practice, number of providers, exposure to telehealth | X |  |  |  |
| Provider practice characteristics | Years in practice, gender, comfort with genetic testing/telehealth/eHealth, champion for genetic testing/telehealth/eHealth | X |  |  | X |
| Patient background | Age, race, ethnicity, education, insurance |  | X |  |  |
| Patient attitude toward genetic testing *and cost of testing* | Attitudes about genetic testing scale<sup>1</sup> |  | X |  |  |
| Patient comfort with technology | Items from the NCI Health Information National Trends Survey including internet and social media use, electronic medical record use, and perceptions of privacy<sup>67</sup> |  | X |  |  |
| Patients baseline genetic knowledge, health literacy | KnowGene Scale,<sup>5</sup> Brief Health Literacy Screen<sup>66</sup> |  | X |  |  |
| Process |  |  |  |  |  |
| Planning | Publicly available digital materials, registration process for local providers, quality of materials describing procedures for patients and providers | X* | -- | X | X |
| Opinion leaders | Which care providers (local providers vs genetic counselors) and insurers are most influential? How do they influence others? What could have helped them to be more effective? |  |  |  | X |
| Champions | Who in the practice/program helps ensure that all steps were completed? What did they do to make the delivery model successful? |  |  | X | X |
| Reflecting and evaluation | What procedures are working (provider registration, provider collaboration with genetic counselor, genetic counselor review of cases for patients in digital arms)? Which are not? What can we change to make the process easier? |  |  | X* | X* |
| * Includes regular personnel and team debriefing about what is working and not working about the digital alternative delivery models |  |  |  |  |  |

## Methods/Design

This study, a **R**andomized Hybrid Type I Effectiveness-Implementation Study of an **e**Health Delivery **A**lternative for **C**ancer Genetic Testing for **H**ereditary Cancer (eREACH2), is a 4-arm randomized study in real-world clinical patients who meet National Comprehensive Cancer Network (NCCN) or American Society of Clinical Oncology (ASCO) guidelines for germline genetic testing to evaluate the effectiveness of an eHealth (e.g. digital) approach compared to the traditional standard of care. We use a non-inferiority study 2×2 design where standard-of-care pre-test (visit 1) and post-test (visit 2: disclosure) counseling delivered by a GC are replaced with an interactive patient-centered digital intervention (**Figure 2)**. We used the SPIRIT (Standard Protocol Items: Recommendations for Interventional Trials) checklist when writing this report.(41)

**Figure 2:**
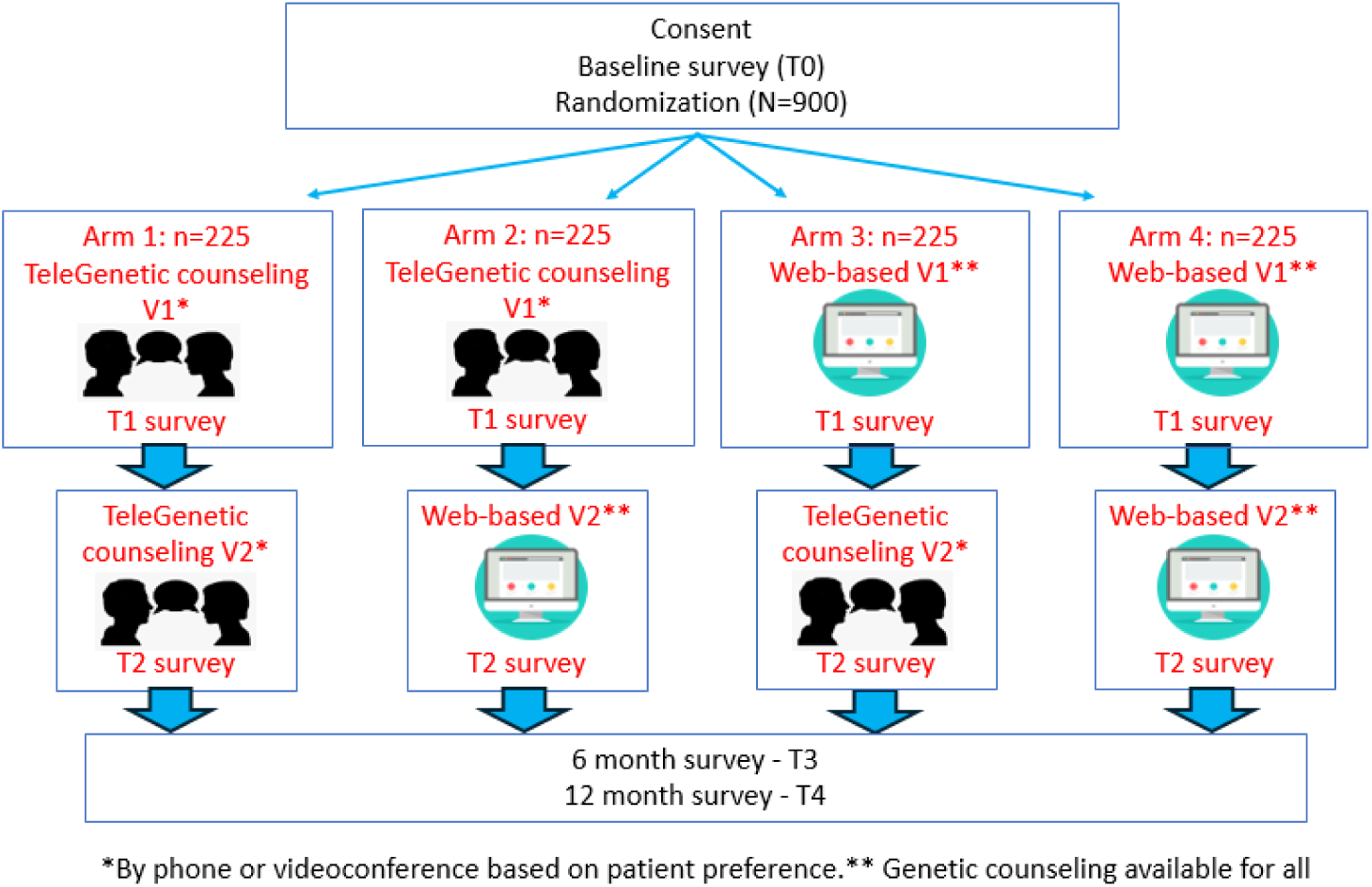
Study Schema. V1: Visit 1 (pretest education; V2: Visit 2 (result disclosure).

The primary aim of the eREACH2 study is to evaluate the short-term effectiveness of offering a digital alternative for pre and/or post-test genetic counseling to provide equal or improved uptake of genetic services and short-term cognitive (e.g. understanding), affective (e.g. distress and uncertainty) and behavioral (risk reducing and screening behaviors and communication to providers and relatives) outcomes. Primary outcomes for Aim 1 include uptake of genetic services, genetic knowledge, and general anxiety. Uptake of testing and all other cognitive, affective, and behavioral outcomes are considered secondary outcomes.

The secondary aim is to assess the effectiveness of digital delivery alternatives to provide equal or improved longitudinal understanding of, reactions to, and use of genetic test results, and to reduce genetic counselor time as compared to the remote traditional two-visit delivery model with a genetic counselor. The implementation aims are to a) understand potential moderators (e.g. intervention usage, sociodemographic factors, genetic test result and test choice) of short-term and longitudinal outcomes to understand who benefits more or less from a digital delivery alternative, and b) facilitators and barriers to implementation eHealth delivery alternatives for genetic services and recommendations for future adaptation and sustainability in clinical practice throughout, and beyond oncology.

The study has been approved by the University of Pennsylvania Institutional Review Board (Protocol# 850242). Any major protocol modifications will be reported in subsequent manuscripts describing study results. Recruitment emails to eligible participants include the study purpose and procedures as well as the direct REDCap (Research Electronic Data Capture; Vanderbilt University) web link to the electronic informed consent form, HIPAA (Health Insurance Portability and Accountability Act) document, and clinical consent to care (for patients new to Penn Medicine). To ensure confidentiality, all responses and case characteristics will be deidentified.

### Patient-centered eHealth Intervention

The patient-centered digital intervention is informed by the tiered-binned model(19), is designed to replace a traditional counseling session with a genetic counselor, and is reported here in accordance with the Genomic Medicine Checklist for reporting on Digital tool development, implementation, and evaluation (GeM-CheckD-**Supplementary Table 1**).(42) The intervention is adapted from the eREACH1 intervention designed specifically for patients with metastatic cancer.(24, 43) The intervention includes Tier 1, indispensable information presented to all users, and optional Tier 2 content (more in-depth information, examples and/or videos). As eligibility includes all testing naive individuals who meet NCCN criteria, educational pathways have been created to customize content based on specific clinical scenarios including personal history of cancer (yes/no), cancer stage, and known familial mutation. Readability testing was completed for all content and modifications made to achieve a readability score of 8^th^ grade or lower when possible.

As shown in **Table 2**, the pre-test intervention includes 8 modules, including 1) Introduction, 2) Purpose of Testing, 3) Genetics Overview, 4) Implications of Results, 5) Possible Results and Implications, 6) Genetic Testing Options, 7) Things to Consider before Genetic Testing, and 8) Testing Decision. Example screen shots are included in **Figure 3**. Completing Tier 1 content takes 11-14.5 minutes to complete for the average user, and 28 min for all content and videos. After viewing the minimum Tier 1 screens, participants are asked to record their decision to either proceed with testing and their testing choice (a targeted testing panel based on personal and family history, a larger panel which can include genes of unclear immediate value, or to speak with a genetic counselor and consider a custom panel to address specific needs or concerns). Participants can also decline testing and are asked to provide their reason. Those who log a decision to move forward with testing have their selection, along with the details of their personal and family history, reviewed by a study genetic counselor. If a study genetic counselor feels an alternative decision for testing may be more appropriate based on family history, participants will receive a call from the genetic counselor to confirm their decision.

**Figure 3:**
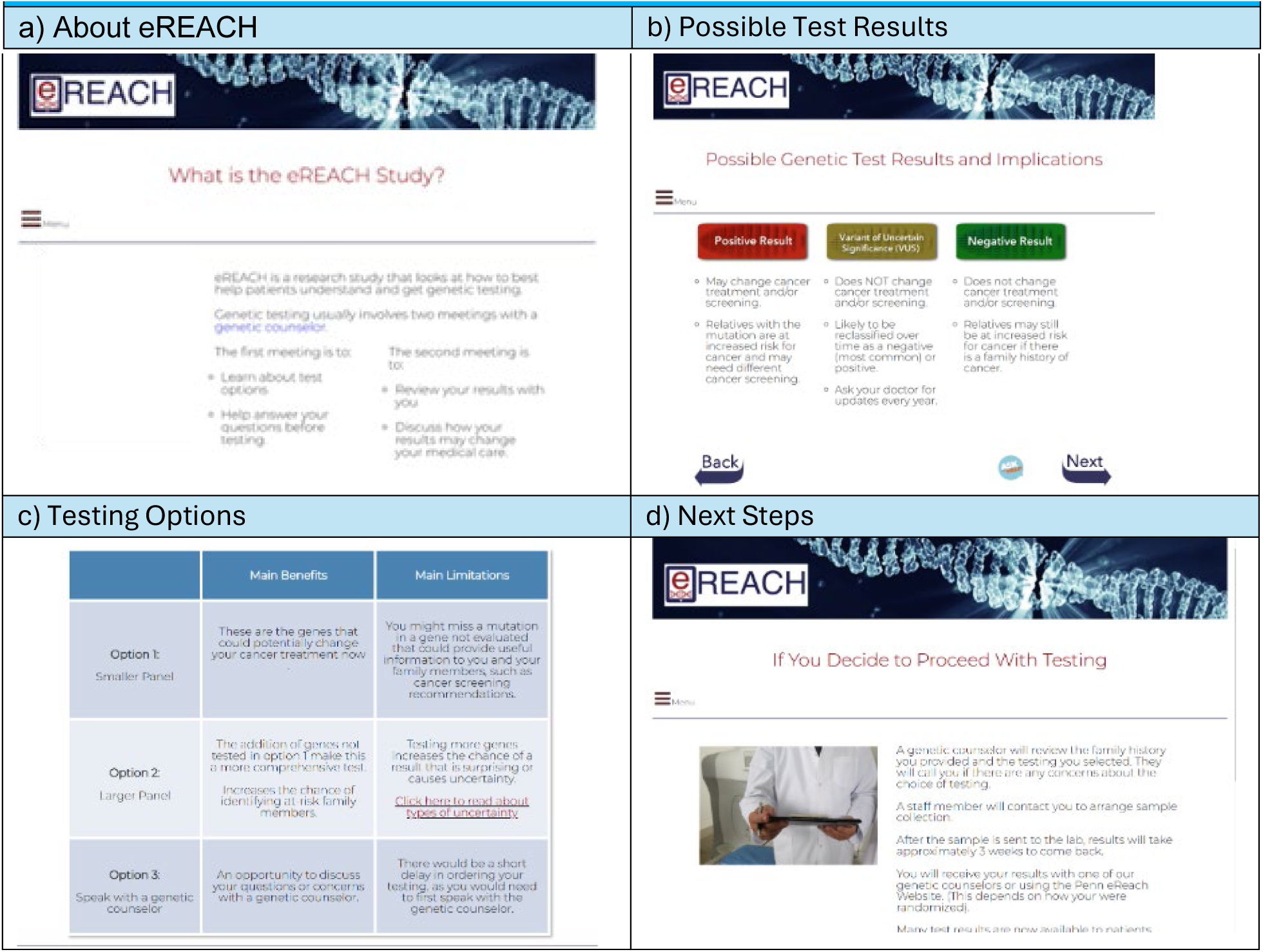
Examples of screens from the pre-test counseling (visit 1) digital intervention

**Table 2:** eREACH2 – Pre-test education intervention*.

| **Module** | **Tier 1 Content\*\* (# screens)** | **Tier 2 Content (# screens)** | **Tier 2 Videos [minutes]** |
| --- | --- | --- | --- |
| WELCOME/LANDING PAGE | NA | NA | NA |
| 1: Introduction | -Introduction (1)<br>-Why have genetic testing? (1)<br>-What is the eReach Study? (1)<br>-How to use this website (1)<br>-What to expect on this website (1) |  |  |
| 2: What is and why genetic testing? | -What is genetic testing? (1)<br>-Why is genetic testing recommended? (2)<br>-How testing might affect cancer treatment or screening (1) | -More information about costs (1)<br>-More information about targeted medications (1)<br>-More information on surgical options (1)<br>-More information on screening and prevention (1) |  |
| 3: Genetics Overview | -Genetics 101 (4 brief sliding screens)<br>- Difference between tumor and inherited genetic changes (1)<br>-How inherited mutations are passed down through families (1)<br>-Difference between sporadic and inherited cancers and BRCA1/2 example (2)<br>-Types of inherited cancer risk genes (e.g. high v moderate risk) (1) | -More information on inherited gene risks (4), including:<br>-- high risk genes (1)<br>--moderate risk genes (1)<br>--limited information risk genes (1)<br>-- risks for cancer with each of the above (1) | Genetic counselor explaining “a brief genetics lesson” [2:10] |
| 4: Implications of results | -How my results might impact my medical care? (1)<br>-How my results might impact my families medical care? (1) | -More information about targeted medications*** (1)^<br>-More information on surgical options (1)^<br>-More information on screening and prevention (1)^ |  |
| 5: Possible results | -Types of results and implications (1) |  |  |
| 6: Genetic Testing Options | -Testing options: small panel, larger panel or speak with a GC (1)<br>-Choose the test that is right for you (2) | - Types of uncertainty (1)<br>-What is a VUS? (1)<br>-How patients choose among the options (1) | Genetic test panel options [3:13]<br>Choosing a custom panel [1:51] |
| 7: Things to consider before genetic testing, including costs | -Benefits, risks and limitations of testing (2)<br>-GINA (1)<br>-Costs of testing (1) | -Types of uncertainty^ (1)<br>--What is a VUS?^ (1) | What is a VUS? [3:47]^ |
| 8: Testing decision and Next steps | -Next steps (1)<br>-Do you have enough information and testing choice (2) | Glossary of terms |  |

\*The intervention is informed by the tiered-binned model for genetic education and informed consent and was reviewed with cancer genetics providers and experts in health disparities and health communication. Readability is 8<sup>th</sup> grade or lower for all screens. Completing Tier 1 content once takes approximately 14 minutes and 27 minutes to complete all content and videos. Videos include a genetic counselor explaining specific topics. Content in some videos are intentionally redundant to Tier 1 and Tier 2 content and designed to provide an alternative method for reviewing content.\*\* There are two different versions (“pathways”). Pathway 1 is for patients with cancer. Pathway 2 is for patients without cancer. All pages offer the “ask for help” button. \*\*\* Only presented for Pathway 1. ^refers to duplicate screens as described above

The patient-centered digital result disclosure (visit 2) intervention includes 4 modules, including 1) Welcome Page and Instructions, 2) Test Result, 3) Explanation of Result, 4) Next Steps (**Table 3**). Completing Tier 1 content takes approximately 4-5 minutes for the average user, and approximately 6-10 minutes for all content and videos. Readability testing was completed for all content and modifications made to achieve a readability score of 8^th^ grade or lower when possible. Example screen shots are included in **Figure 4**.

**Figure 4:**
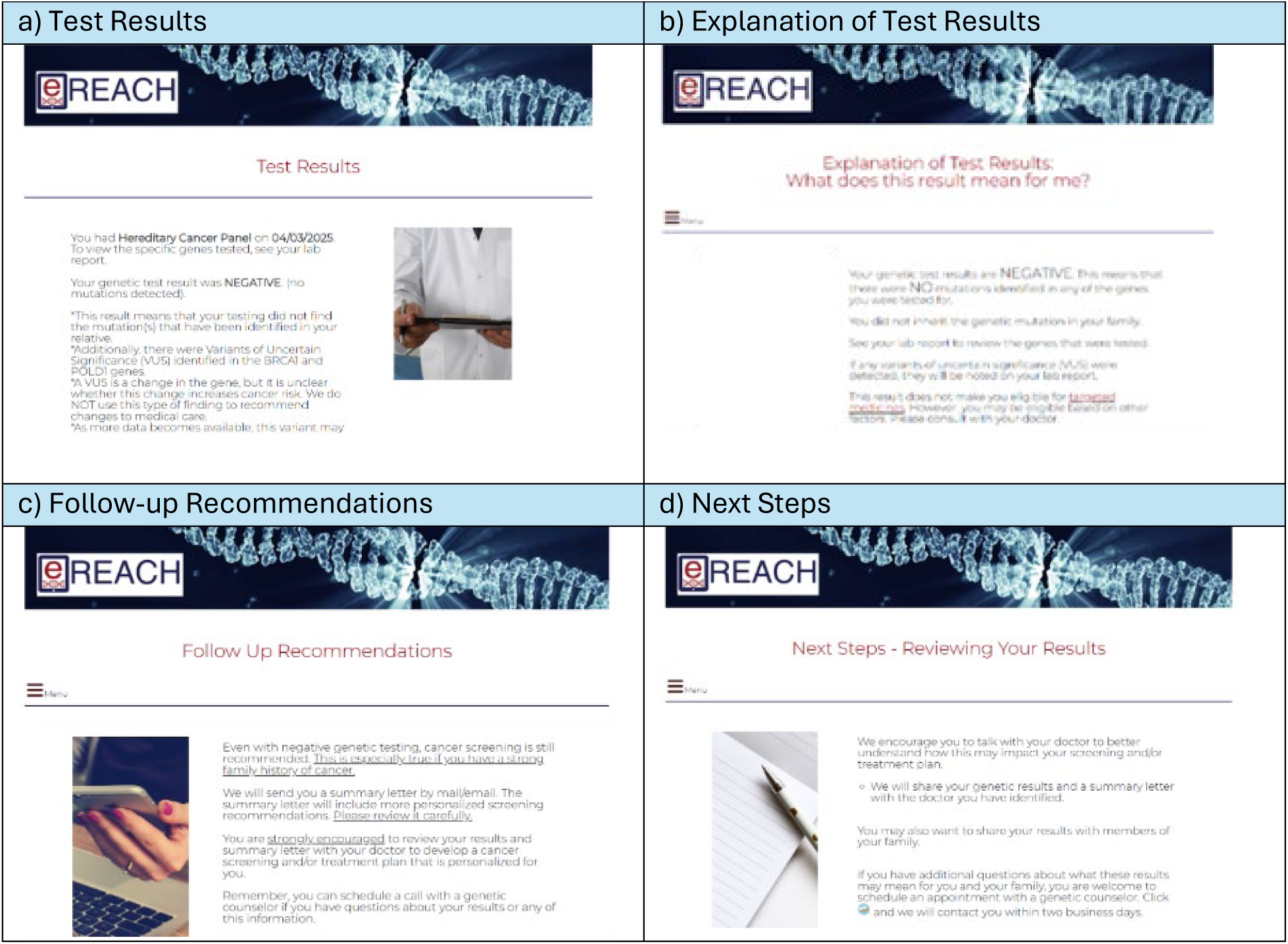
Examples of screens from the post-test counseling (visit 2) digital intervention

**Table 3:** eREACH2– Test disclosure intervention.

| **Table 3: eREACH2– Test disclosure intervention** |  |  |
| --- | --- | --- |
| **Module** | **Tier 1 Content\*\* (# screens)** | **Tier 2 Content (# screens)** |
| 1: Welcome Page/<br>Instructions | -Introduction, navigation instructions including how to ask for help (1) | -Option to review select educational material (5)<br>--Why were you referred? (1)^<br>--Genetics 101 (1)^<br>--Targeted medications/screening (1)^<br>--Somatic v. inherited mutations (1)^<br>--What is a VUS (1)^ |
| 2: Test Result | -Confirm readiness (1)<br>-Genetic test result, summary of genes tested (1) | -Difference between tumor (e.g. somatic) and inherited genetic changes (1) |
| 3: Explanation of Result | - Explanation of result (1)<br>-What does this result mean for me? (1)<br>- Follow-up recommendations (1)<br>-What does this mean for my relatives? (1)<br>-FOR POSITIVES ONLY – General medical management recommendations (1) | For VUS results only:<br>-What is a VUS?^ (1)<br>--What is a VUS? [3:47]^ |
| 4: Next Steps | -Option to speak with a GC if you have questions and recommendation to f/u with your doctor (1)<br>- Recommendation to share results family members (1)<br>- How to download and read your test report (1) | -PDF of clinical test result available for download |
| Completing Tier 1 content once takes approximately 4.5 minutes and 6 minutes to complete all content and videos.<br>^duplicate screens from Pre-test Education |  |  |

### Design and Development of Digital Intervention

The initial intervention content was reviewed with 13 patients purposively selected to represent a range of ages, gender, education and cancer history through individual user testing interviews (**Table 4**). We included participants who previously had and did not have genetic counseling and testing. Participants were asked to comment on the clarity, usefulness and appearance of each screen. They were also asked to comment on proposed video topics, including usefulness, what they would expect to see covered in the video, and the likelihood that they would watch the video. Interviews were reviewed to identify recommendations for changes or areas of confusion. Two research team members reviewed and selected key comments for consideration and at least two members and a GC reviewed comments to consider changes to the content. All specific recommendations were considered, and particularly those with more than one participant recommending a similar change. In scenarios where user testing participants had conflicting recommendations, the team used best judgement to make changes. The most common recommendations for both visit 1 and visit 2 interventions were to simplify content (e.g. use bullets, chunk information), address confusing content and terms, emphasize benefits to testing, move some content to Tier 2 and address costs of testing earlier (**Supplementary Table 2**).

**Table 4:**
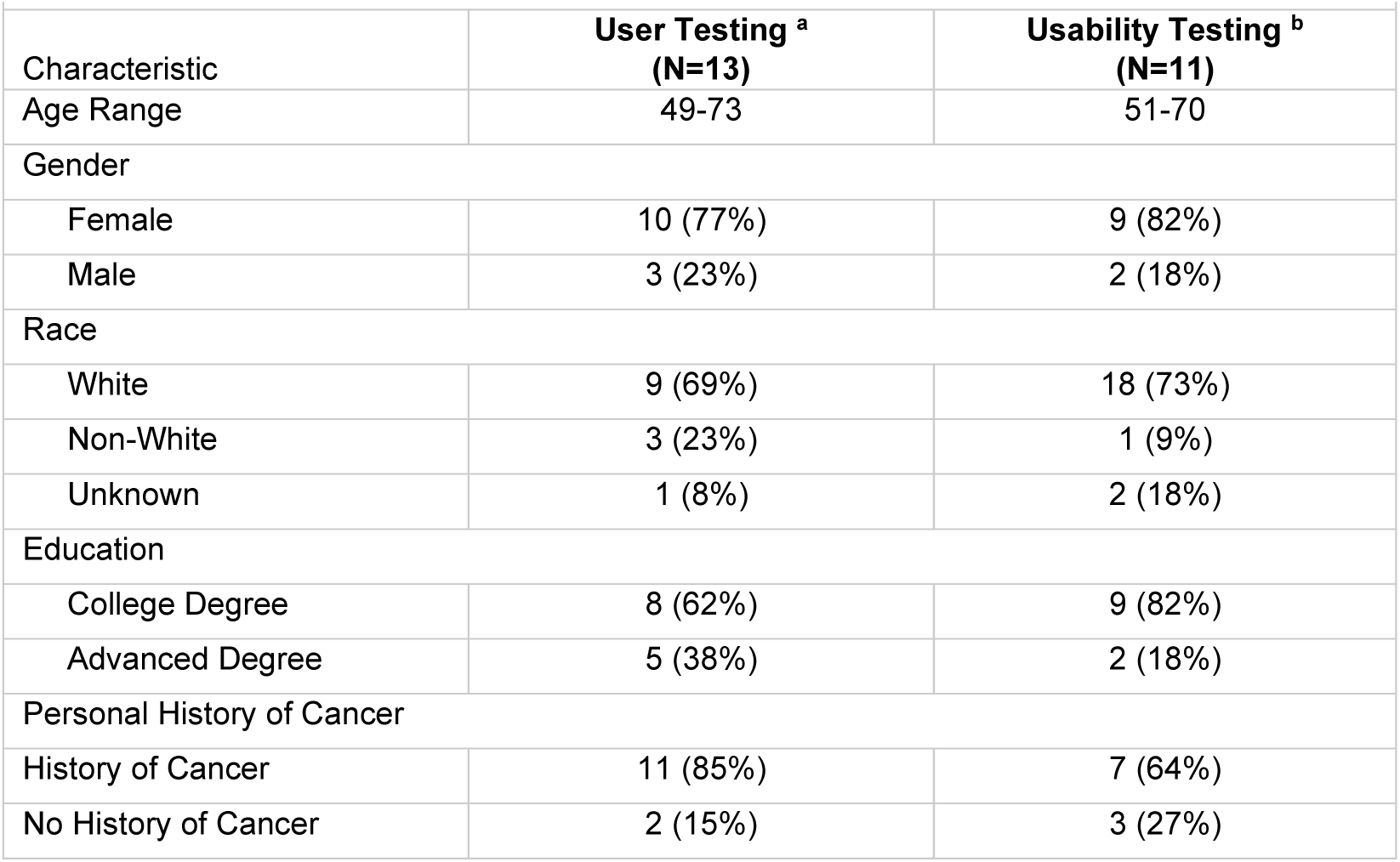
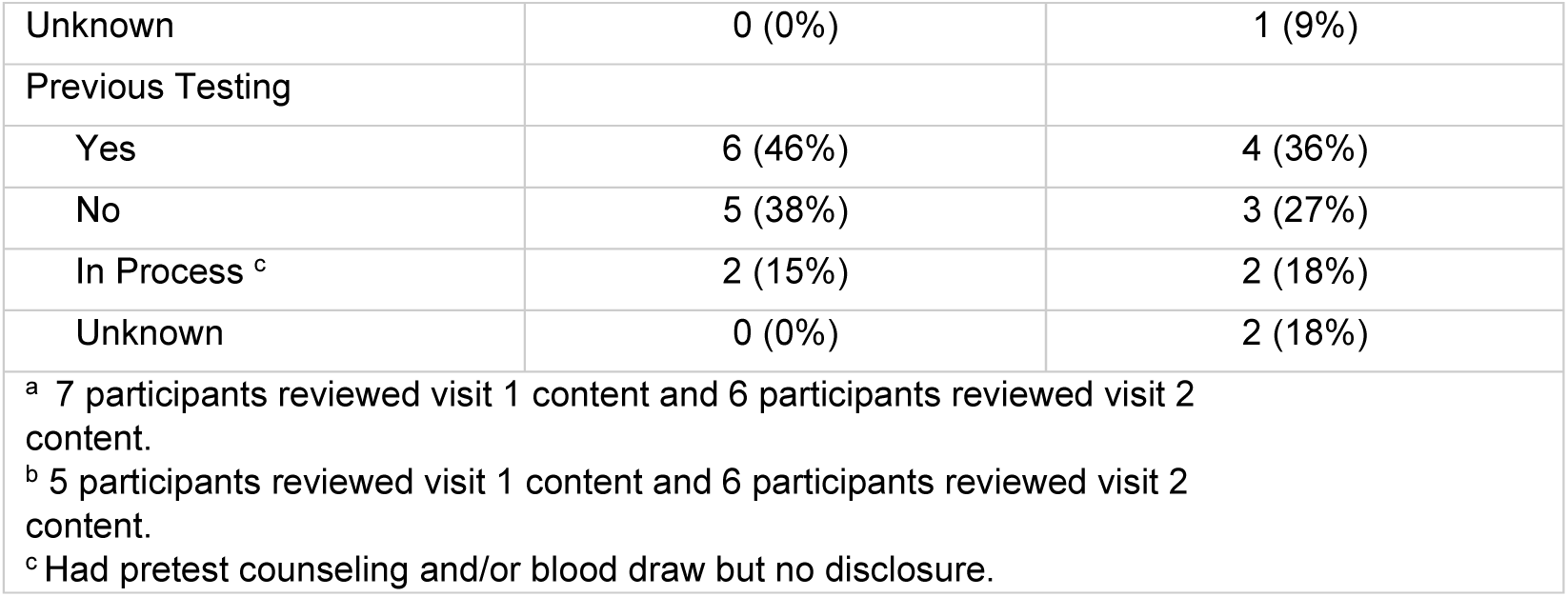
Characteristics of participants who completed eREACH2 User and Usability Testing.

We completed usability testing with 11 participants (**Table 4**). These participants used the digital intervention and were asked to comment on things that were confusing, functions that were challenging, and other changes they suggest. A research assistant viewed their navigation through co-browsing software. Several function challenges were identified, prompting revisions to the intervention (e.g. labeling or size of arrows). Participants also commented on the size of text and images and had recommendations regarding photos (**Supplementary Table 2**).

### Setting

This research is conducted at the University of Pennsylvania, but seeks to recruit a real-world and representative patient population, with an emphasis on patients with access barriers to genetic testing, including patients in community practice settings, where access to genetic counselors is more limited.

Remote genetic counseling services in this study are provided through the Penn Telegenetics Program, which has developed procedures for individual patients and providers across the nation to access remote genetic counseling services. For patients outside Pennsylvania, the Penn Telegenetics counselors collaborate with local health care providers to provide genetic services in the home. This model includes a physician registration process to facilitate this collaborative care model and has been successfully utilized in related nationally accruing studies (NCT04455698, NCT05427240). The Penn Telegenetics genetic counselors are licensed in all US States as required by state licensure laws.(44)

### Study Participants

#### Eligibility

Eligibility criteria include individuals who are 18 years of age or older, able to understand and communicate in English, have had no prior clinical germline genetic testing, and meet current NCCN or ASCO guidelines for germline genetic testing. Exclusion criteria include uncorrected or uncompensated speech, vision or hearing defects that would lead to the participant being unable to communicate effectively with a medical provider, uncontrolled psychiatric/mental condition or severe physical, neurological, or cognitive deficits rendering the individual unable to understand study goals or tasks.

We utilized digital tools to collect family and personal cancer history to assess eligibility according to clinical guidelines at the time of enrollment (i.e. NCCN and ASCO). This initially included a commercially available chatbot, although we transitioned to a REDCap survey in March 2024. Participants can also complete the family and personal history questions with a research staff member. Research staff and GCs assess responses to determine if patients meet guidelines for testing.

#### Recruitment

Eligible patients can be provider-or self-referred from a range of recruitment sources. 1) The Penn Telegenetics program provides remote clinical services at several regional centers without GCs on site. Clinical coordinators and providers at these sites identify eligible patients in their practices and refer them to the research team. 2) Social media campaigns through advocacy partners including Cancer specific support groups (e.g. FORCE-Facing Hereditary Cancer Empowered, Cancer Support Community) advertise the eREACH2 study through study flyers, website banners, etc. Patients can contact the research team and enter the study as self-referrals. In collaboration with the University of Pennsylvania Office of Clinical Research, the team posts Facebook ads on ClinicalTrials@Penn directed towards patients and providers highlighting the utility of genetic testing in various populations. Interested patients can enter the study as self-referrals. 3) Patients within the Penn Network with a long wait for services or urgent indication (e.g. need genetic testing quickly for a medical or surgical decision) are offered eREACH2 as an alternative. 4) eREACH2 study information has been shared with patients identified with a genetic mutation through the UPenn Cancer Risk Evaluation Program and Penn Telegenetics Program to share with their relatives who are candidates for cascade testing live outside our catchment area. Potential participants receive up to five contacts, typically three emails and two phone calls. Recruitment invitations began in November 2022.

#### Enrollment Goals

Our enrollment goal for the primary aim is 203 patients per arm. To achieve this goal, we seek to recruit up to 1000 patients (250 assigned to each arm), prioritizing those who have access barriers to genetic testing.

### Study arms

Once participant eligibility is determined using the personal/family history collected, and have completed informed consent with a study coordinator and the baseline (T0) survey, they are randomized into one of the four study arms (**Figure 2**). Randomization is stratified by gender, using a permuted block design within strata for randomization. The allocation sequence was generated by the study biostatistician and made available to research staff. There was no blinding after randomization.

#### Pre-test counseling (Visit 1)

Participants randomized to traditional genetic counseling (Arms A and B) complete visit 1 by phone or videoconference (according to patient preference) in the location of their preference. Participants who select videoconference are provided links to download secure videoconferencing software on their home computer or device. The Penn Telegenetics team utilizes a HIPAA-compliant technology platform for videoconference visits. In our communication protocols, if videoconference technology fails, GCs convert the session to phone. To assess fidelity, all visits are recorded unless requested otherwise by the participant. Additionally, a clinically available Visual Aid Packet is recommended for all participants receiving their V1 standard-of-care counseling by telephone or videoconferencing, consistent with clinical practice for remote counseling. Penn Telegenetics counselors utilize communication protocols adapted from related studies.(28, 45) The Telegenetics team contacts participants up to 5 times to schedule their pre-test counseling session. If a scheduled participant does not show for their appointment, they are contacted 3 times to reschedule. If Visit 1 is not completed, then the respective provider is notified (if participant was provider-referred).

Participants randomized to the digital intervention (Arms C and D) are provided instructions for obtaining a private code to access the pre-test counseling digital intervention. Participants are informed that they can choose to schedule a counseling session with a GC if they do not wish to access the digital intervention or do not have online access. They can also ask to speak with a GC at any point. Up to 3 electronic reminders and 2 telephone reminders are sent by members of the research team to participants randomized to this arm who do not access the digital intervention.

Participants are asked to complete a post-visit 1 survey (T1) within 72 hours (3 days) of completing visit 1, though surveys which are completed up to 7 days after are still accepted.

#### Genetic Testing

In all study arms, there are 3 available options for testing (**Figure 5**).(43) Option 1: a smaller panel based on personal and family history and focused on genes that have strong evidence to change cancer care or screening. Option 2: a larger or more comprehensive panel which could include genes of unclear immediate value but that could have a potential impact on cancer treatment or screening in the future. Option 3: A customized panel based on other health concerns or a desire to exclude particular genes. For Option 3, participants are asked to speak with a GC to review their concerns and options. Those who log a decision on the digital intervention (Arms C and D) have their selection, along with the details of their personal and family history, reviewed by their assigned study GC. In the event the counselor feels an alternative decision for testing may be more appropriate (i.e., participant selects the smaller targeted panel when a large or custom Panel may be indicated), the GC calls the participant to confirm their decision.

**Figure 5:**
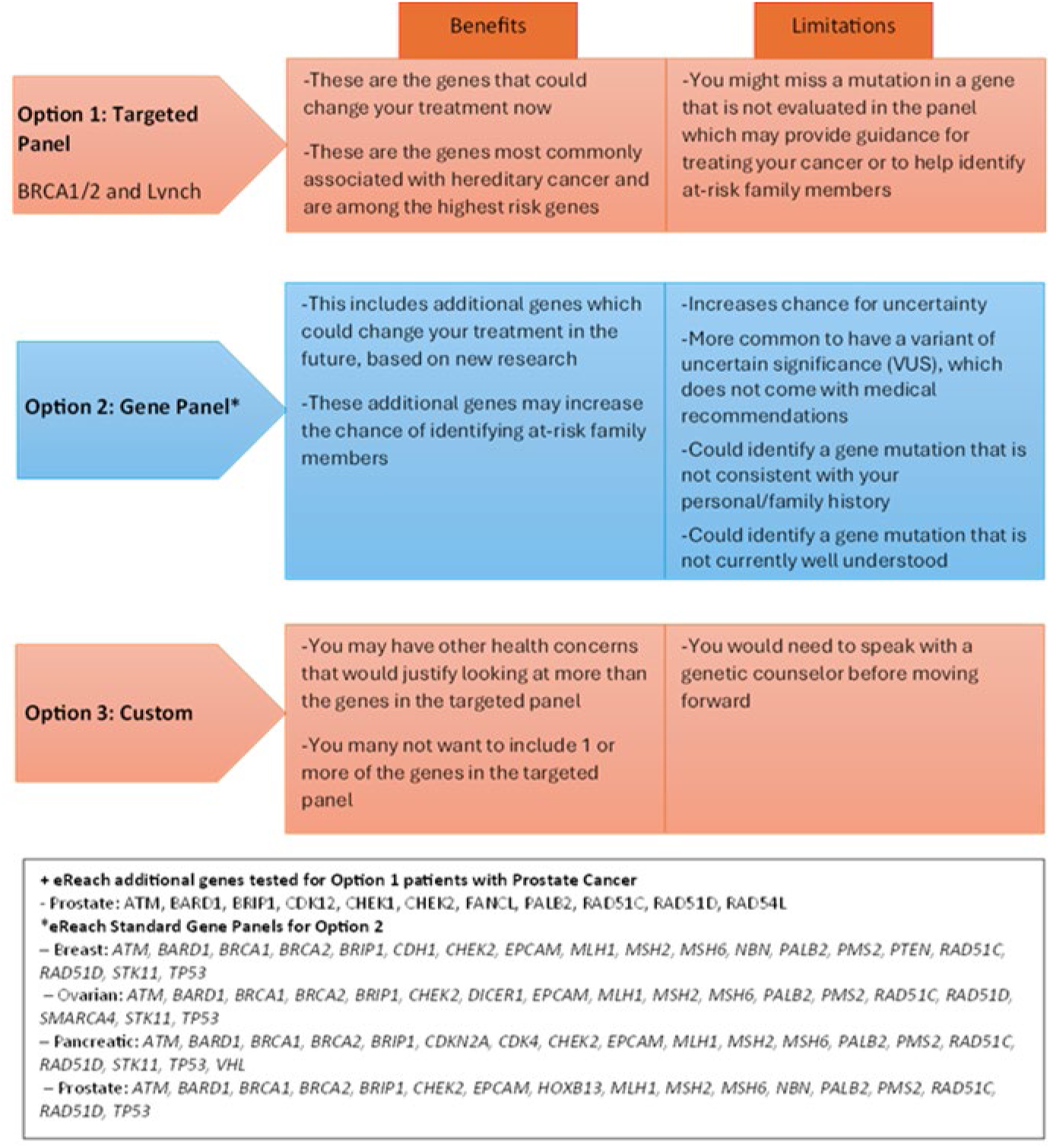
Participant options for genetic testing

Self-referred participants are asked to provide a health care provider’s contact information, to serve as the ordering provider with the GC as required by state licensure laws. The Penn Telegenetics clinical team contacts the provider’s office to provide information about the program and procedures and confirm the provider’s information to include on the testing order. Consistent with current clinical care, all genetic testing is sent to a commercial laboratory. The genetics laboratory conducts an insurance benefits investigation for participants and if there is any out-of-pocket cost >$100, the laboratory contacts the participant. Patients are instructed to contact the GC for any questions regarding coverage and cost. Test kits are sent to the participant’s home, and participants get up to 3 reminders to return their test kits.

#### Disclosure of results (Visit 2)

Those randomized to genetic counseling for Visit 2 (Arms A and C) are scheduled for a telephone or videoconference session with a GC to share their results. The GCs complete a summary note sent to the patient and provider, complete disclosure checklists, and record sessions for fidelity. Staff members check in with participants randomized to this arm who do not schedule an appointment with a GC by email and/or telephone up to 6 times. If staff are unable to contact the participant, the registered/ordering provider will be notified.

Those randomized to the digital intervention for Visit 2 (Arms B and D), are provided with their result and gene specific screens (as appropriate) on the private intervention website. The results are entered and reviewed by two study team members (research member and GC) for accuracy and quality control. Participants are notified by email that results are available and can be accessed, and that they can request to speak with a GC if they prefer, before or after viewing their results. After return of results, the GC provides the referring or registered provider with the genetic test results and chart note. GCs are available to answer any patient or provider questions. Up to 3 electronic reminders, coupled with 2 telephone reminders are sent to participants randomized to this arm who do not access the digital intervention.

### Outcomes

#### Uptake of testing and Patient-reported Outcomes)

Theoretically informed patient-reported cognitive, affective and behavioral outcomes of genetic services are collected at baseline (T0), after Visit 1 (T1) and Visit 2 (T2) and at 6 and 12 months (T3, T4).

*1) Uptake of counseling and testing (primary non-inferiority outcome)* is assessed through Penn Telegenetics service records for those assigned to (or who select for) standard-of-care genetic counseling. Uptake for those assigned to the digital intervention are recorded as completion of the digital intervention.
*2) Knowledge of genetic disease (primary non-inferiority outcome)* is evaluated at all time points (T0-T4) using the KnowGene Scale, a 16-item scale administered to patients after genetic counseling and testing to measure their understanding of the health implications of genetic test results.(46) It includes constructs such as health implications to oneself as well as relatives and covers penetrance, actionability, limitations of current technology, and monogenic inheritance patterns and is scored from 0 to 16 (correct answer = 1; incorrect/don’t know response = 0) with higher scores representing greater genetic knowledge.(46) Perceptions of genetic disease and risk reduction options will be evaluated using a series of scales which include perceived risk (3 items), perceived controllability (10 items), timeline (1 item), and utility (6 items) are collected at T0, T2-T4.(31) Recall of result is collected at T2-T4.(47) Collectively, these items will provide an opportunity to assess for individual misunderstanding (e.g. incorrect test result recall, changes in perceived risk that don’t align with test results).
*3) Reactions to genetic information* are assessed with multiple instruments that evaluate psychological distress, adjustment, and satisfaction. a) *General anxiety (primary non-inferiority outcome) and depression* are assessed by the respective 4-item short Patient Reported Outcomes Measurement Information System (PROMIS) scales.(48) b) *Disease-specific distress* is measured using the 8–item Impact of Events Scale (IES) (alpha = 0.82-0.90),(49–52) consistent with a growing body of literature identifying a broader range of cancer-related stress reactions including intrusive thoughts and feelings and avoidance in response to risk of cancer.(53–56) c) *Satisfaction with genetic services* is assessed at T1 and T2 with a 14-item scale previously used in other studies evaluating alternative genetic testing delivery models (alpha = 0.73-0.88).(16, 24, 57) d) *<u>Psychosocial impact of genetic testing</u>*, including positive responses and uncertainty are assessed using the Multi-dimensional Impact of Cancer Risk Assessment Questionnaire (MICRA) at T2-T4. The MICRA has been utilized in many genetic studies to evaluate distress, uncertainty and positive responses to receipt of genetic test results and is well-suited to evaluate change over time.^58^ e) *Decisional regret* is assessed at T2-T4 with the 5-item validated Decision Regret Scale (Cronbach α = 0.81-0.92).(58, 59) f) Informed decision making is assessed with the Attitudes Toward Genetic Testing (ATGT) scale at T1 (Cronbach *α* = 0.73),(60) knowledge (T1) and decisions to proceed with testing, as previously utilized in our studies and others.(19, 60, 61)
*4) Use of genomic information* includes treatment change based on genetic test results (for affected participants, T3-T4), behavior change at T0, T3, T4 (e.g., exercise, tobacco and alcohol use), performance of risk reduction and cancer screening behaviors (T0, T3-T4), and intent to communicate (T2-T4) to at-risk relatives and health care providers.(16, 62)
*5) Provider time* is recorded per patient for a subset of participants to estimate provider time by arm. This includes those in Arms 2-4, where services have the potential to be completed partially or entirely by the digital intervention. These patients still have a GC review their history, testing decision, support testing submission, review results and the disclosure information provided, and be available for questions upon request. Thus, in all arms, GC time for case review, genetic counseling, and conversations with patients and their providers are recorded.

### Moderators of Uptake and Patient Reported Outcomes

Moderators are collected at baseline and include sociodemographics, health literacy, and comfort with technology. Sociodemographic data includes race/ethnicity, education, marital status, gender, age, zip code, health insurance status, usual source of care status, employment status, financial toxicity^65^ and household income. History of cancer and genetic test results (positive, negative, true negative and type of testing (targeted versus panel)) are obtained from Penn Telegenetic Program records. Participants are asked at T2 about any out-of-pocket costs for genetic testing. Health literacy is assessed at baseline (T0) with the Brief Health Literacy Screen (BHLS) which has been validated to detect inadequate health literacy in clinical medical populations^66^. Comfort with technology is assessed with selected items from the NCI Health Information National Trends Survey (HINTS), including internet and social media use electronic medical record use and perceptions of privacy (7 items).^67^

Additionally, individual use of the digital intervention is recorded throughout the study to evaluate the impact of intervention usage (e.g. dosage) on outcomes and who benefits more and less from the intervention. Intervention use includes actual usage, adherence and attrition. For actual usage, we record the time each participant logs in and logs out. Actual usage is evaluated for the intervention in general and secondarily for specific intervention content (e.g. videos, specific pages). Adherence is defined as completing the full Tier 1 content at least once. Rate of attrition will be defined as the number of days from the end of the first week to the day when no login or activity has occurred for over 90 days.

### Implementation Outcomes

CFIR-informed constructs have been selected to evaluate factors related to uptake of genetic services, and implementation facilitators, barriers and adaptations (**Table 1**). We will collect additional quantitative and qualitative data from multiple key informants including:

1) Patients: A purposive sample of 15-30 patients will be selected to maximize representativeness in terms of gender, race/ethnicity, education and uptake (enrolled but did not complete counseling, completed counseling and completed testing).
2) Local providers: 15-30 local health care providers identified by participants to collaborate for delivery of remote telegenetic services. As above, we will seek to maximize representativeness in terms of gender, race/ethnicity, years in practice, type of practice and uptake (registered with patient and completed services, registered but did not complete services, declined registration);
3) Genetic Counselors: 15-30 GCs not participating in the study will be identified to provide feedback on implementation of the digital interventions and remote telehealth services in their field and practice. As above, we will seek to maximize representativeness in terms of gender, race/ethnicity, years in practice and type of practice.
4) Insurers: 5-8 individuals working in health care coverage and insurance will be identified to provide feedback on coverage of digital and remote telehealth services.

We will use the CFIR interview guide (63) to develop study-specific open-ended items covering CFIR constructs as outlined in Table 4. We seek to continue recruitment to the above enrollment goals or until data saturation occurs across the theoretical domains (e.g. no new themes introduced within the constructs included in key informant interviews).(64)

## Data Analysis Plan

### Primary Non-inferiority Analysis

We hypothesize that the intervention arms incorporating the digital delivery alternatives, with access to a genetic provider upon request, can provide equal or improved short-term and longitudinal cognitive, affective and behavioral outcomes and lower provider time. Our primary outcomes are 1) uptake of services defined as completion of testing or completion of visit 1 (to account for patients who make an informed choice to decline testing after pre-test education/counseling), and change (baseline to T2) in 2) knowledge and 3) anxiety. We will test whether digital delivery alternatives are non-inferior (knowledge and anxiety) or equivalent (uptake) to traditional counseling. In non-inferiority and equivalency tests, the null and alternative hypotheses are the reverse of usual; for non-inferiority, the null hypothesis is that the digital delivery alternative is worse than the traditional delivery model. We will conclude that the digital delivery alternative is non-inferior using a modified non-inferiority ANOVA test (knowledge and anxiety) or a modified equivalency Chi-squared test (uptake). We will use an intention to treat approach, comparing randomization arms, but we will also confirm results in secondary analyses with per-protocol analyses; many recommend that both intention-to-treat and per-protocol analyses be consistent to declare non-inferiority or equivalency.(65)

In secondary analyses, we will test pairwise differences among the four groups using T-tests (knowledge and anxiety) and chi-squared tests (uptake). We will also examine the balance of potentially confounding baseline covariates among the arms. If there are covariates that vary significantly among the arms, we will estimate effects using multiple linear or logistic regressions in which we include indicator variables for randomization arms as well as the potential confounding covariates. We will repeat all analyses for the secondary outcomes using similar methods, including assessment of individual misunderstanding of results (e.g. incorrect test result recall, changes in perceived risk that don’t align with test results).

As treatment effects might differ based on enactment of the Cures act (pre-versus post-implementation of immediate genetic test release to patients at the University of Pennsylvania), we will estimate interaction regression models in which we include binary (0/1 no/ yes) time period (pre-July 2023 versus post-July 2023), randomization arm, and the interaction between time period and randomization arm (randomization arm indicators times [x] time period) as covariates in the model. In addition, we will examine interaction models in which we compare outcomes of people who receive results prior to their study-protocol disclosure method appointment with those who do not receive results prior to their study-protocol disclosure method appointment. We will exclude those who do not receive results at all in this secondary analysis examining the potential moderation effect of pre-appointment disclosure.

#### Power justification (Aim 1)

We chose our 3 primary variables since they demonstrated possible change between groups in our preliminary work, the Returning Genetic Research Panel Results for Breast Cancer Susceptibility (RESPECT) study.(62) We will use a non-inferiority test (knowledge and anxiety) or an equivalency test (uptake), and to be conservative will require non-inferiority or equivalency in both the intention-to-treat and per-protocol analyses, as recommended. (66) We will use a Bonferroni correction assuming three hypothesis tests to set the nominal level of statistical significance (0.05/3=0.0167 nominal p-value).

Our non-inferiority test for the two continuous variables will be based on an ANOVA F-statistic test jointly comparing the four randomization arms. We will fail to reject the null hypothesis of inferiority if the joint p-value is less than 0.2 *and* all of the three experimental web arms have a standardized effect of 0.138 standard deviation units or worse (with the direction standardized such that higher values imply beneficial change) when compared to the control GC only arm. This design will approximate a one-sided test, so that we will declare non-inferiority if the intervention arms have better outcomes. We chose a 0.138 cut-point because it is a small standardized effect. With 203 participants per arm, we have approximately 1.67% non-inferiority Type I error and 88% to 90% non-inferiority power (see **Table 5**). In calculating power, we assumed that the four change score standardized means (standardized to have change score standard deviation one within group) relative to the control group were 0 (control group reference change),-0.2 (single web intervention arms), and-0.39 (double web intervention arm). These were conservative estimates based on standardized RESPECT web intervention findings of 0.3 change score improvement in anxiety and 0.4 change score improvement in knowledge. We hence have excellent non-inferiority power. The alternative hypothesis for the power calculations was that all arms had equivalent change scores (three intervention arms had 0 mean change relative to the reference control group).

**Table 5:** Sample size needed under various null assumptions about change in continuous variables relative to the control arm, and differences in uptake among arms. Non-inferiority 1-sided Type-1 error or equivalency 2-sided Type-1 error=1.67%, **power ≥ 88%**.

|  |  |  |  |  |  |
| --- | --- | --- | --- | --- | --- |
| Interpretation | Reference Change Arm A | Mean Relative Change Arms B & C | Mean Relative Change Arm D | Absolute Effect Size = SD of means | Approx. N per Arm |
| Worse change in 3 arms. | 0 (reference) | -.22 | -.38 | 0.135 | 215 |
| Worse change in 3 arms. | 0 (reference) | -.20 | -.39 | 0.138 | 203 |
| Worse change in 3 arms. | 0 (reference) | -.24 | -.42 | 0.149 | 175 |
|  | % Uptake Arm A | % Uptake Arms B & C | % Uptake Arm D |  | Approx. N per Arm |
| Worse uptake in 3 arms. | 84% | 75% | 66% |  | 215 |
| Worse uptake in 3 arms. | 84% | 75% | 65% |  | 203 |
| Worse uptake in 3 arms. | 85% | 75% | 65% |  | 175 |
| SD=standard deviation |  |  |  |  |  |

For the uptake rate, the primary test will be an equivalency test rather than non-inferiority. If equivalency is rejected, then we will examine the pairwise direction of effects in secondary analyses, as described in the next paragraph. We will conduct a Chi-squared test of the 4×2 table of uptake among the four arms, but require a p-value of 0.1 or greater to declare equivalence. Assuming uptake proportions of 84% in the traditional care control group (Arm A), 75% in the single eHealth delivery arms (Arms B and C), and 65% in the combined eHealth delivery arm (Arm D), we would need approximately 203 per arm to have similar Type I error and power as above.

Our tests for non-inferiority or equivalency may not fully characterize the findings if there are heterogeneous qualitative effects among the four arms, such as the intervention being beneficial in some arms, but harmful in others. Hence, in secondary analyses, we will examine pairwise comparisons between arms using T-tests (continuous outcomes) and Chi-squared tests (binary uptake outcome). We will perform one-sided tests of the binary uptake outcome to determine the direction of the effects if equivalence is rejected. As our three intervention arms are so similar, we do not anticipate such heterogeneous effects among the intervention arms.

Power calculations were made by PASS 11 in conjunction with R simulations.

#### Analyses for Aim 2

We hypothesize that the intervention arms incorporating the eHealth delivery intervention, with access to a genetic provider upon request, can provide equal or improved longitudinal cognitive (e.g. understanding), affective (e.g. distress and uncertainty) and behavioral (risk reducing and screening behaviors and communication to providers and relatives) and lower collective GC time. As secondary longitudinal analyses, we will perform non-inferiority analyses of the T3 and T4 outcomes using the same non-inferiority criteria as described above. We will also examine time trajectories using regressions estimated by Generalized Estimating Equations (GEE) to account for within subject correlation over time. We will include main and interaction terms of randomization arm with time panel indicators. In tertiary moderator (i.e. effect modifier) analyses, we will include potential confounding variables in the models, as well as all 2-way interactions of time, randomization arm, and confounding covariates. We will also examine 3-way interactions of time, arm and confounders separately for each confounder. Interactions (2-way) are created by multiplying 2 variables together; 3-way interactions are created by multiplying 3 variables.(67)

#### Intention to treat versus other analyses

While the primary analyses will use an intention-to-treat paradigm, we will repeat all analyses but assign participants to per-protocol (i.e. restricting sample to those who properly used the method of pre-/post-test delivery as assigned) or as-treated groups (e.g. assigning all participants to a group based on the method of pre-/post-test delivery used, regardless of group assignment).

#### Missing Data

We will use the multiple imputation methods of Raghunathun et al. for the primary analyses of interest.(68) We have used this method in our related studies, including the randomized COGENT study.(16, 21) Current recommendations for using multiple imputation in the context of non-inferiority trials are ambiguous as the issue is still being researched.(69) Hence, we will contrast our findings with the complete case analyses in sensitivity analyses.

#### Mixed-methods CFIR-informed implementation analysis

We hypothesize that our CFIR-informed multi-stakeholder mixed-methods process evaluation will identify barriers to uptake and inform recommendations for future adaptation and sustainability. Qualitative data will be analyzed using an iterative, deductive content analysis approach and CFIR as the coding framework.(37, 40, 70) The qualitative data will include: a) key informant interviews; b) notes and debriefing from our Telegenetics service and research team meetings. Guided by consensual qualitative methods(71, 72), two independent coders will review notes and assign responses to CFIR constructs. The two coders and qualitative researchers will meet to discuss and reach consensus on any coding discrepancies. Once the coding is complete for the qualitative items, we will triangulate the qualitative and quantitative data (from patient surveys and physician registration), using a convergent mixed-methods approach.(73) The quantitative and qualitative data will be organized by the 5 CFIR domains to evaluate which constructs were associated with, or more prevalent among: a) patients who had testing, testing choices (e.g. targeted testing v. panel v. other options) and those who did not (test yes v. test no), b) patients who completed eHealth alternatives versus those who requested a GC, and c) local providers who registered and their patient completed testing vs. local providers who required multiple requests to register or never registered, providing an opportunity to comprehensively evaluate facilitators and barriers to implementation of remote Telegenetic and digital delivery alternatives to make recommendations for future adaptation and sustainability beyond this study. We will use descriptive statistics, correlations, and generalized linear models, assuming appropriate link and family functions, to examine quantitative variables associated with barriers and uptake and triangulating with the qualitative data. Where necessary, we will estimate models by Generalized Estimating Equations to account for correlated data.

## DISCUSSION

We hypothesize that our digital delivery of pre-test and/or post-test genetic counseling will provide equivalent or improved outcomes compared to the traditional two-visit standard of care delivery model with a GC. Results from this trial will be presented at national conferences and published in peer-reviewed journals. This model has the potential to expand access to genetic services by leveraging digital technologies. Additionally, a remote telegenetic services model, conducted in collaboration with local medical providers, will reach patients across the nation, and in areas of low access. If effective, this model could inform evidence-based guidelines for delivery of medical genetic testing and is broadly applicable and adaptable for other populations and settings even beyond oncology.

### Limitations and Potential Challenges

We acknowledge several potential challenges. Some participants may not have web-access, although we expect this will not be a barrier for most participants given data that 90% of households have broadband internet including 87% of rural households and 96% of adults in the United States report using the internet.(74, 75) Further, understanding how frequently internet access is a barrier to delivery will be critical to understanding how to equitably implement digital tools in health care. Other social determinants of health such as education, financial stability, and experiences with discrimination could also be barriers to participation, but we still expect to be able to recruit a diverse sample because of our wide-ranging recruitment methods from provider referrals to social media campaign.

While our preliminary data and related studies support interest in digital delivery alternatives,(*22, 76*) interest in using a digital tool instead of directly communicating with a health care provider, could be lower than expected. Even if lower than expected, this study will provide critical data regarding patient preferences and required provider resources for implementation. Further, even if only a subset of patients are comfortable with digital delivery, this will likely still reduce overall provider time and increase provider availability and patient access. Limitations in phone and videoconference counseling reimbursement could limit the dissemination of digital delivery models. While these present current barriers, we expect increasing evidence of telehealth benefits will provide strong rationale for appropriate coverage in the future. Patients with lower health literacy may be at greater risk of unfavorable outcomes with digital tools. We attempted to ensure a representative sample of participants in our user and usability testing and in design of our tools. Vulnerable populations are already at risk of not receiving effective communication within traditional health care environments at baseline, so it is also possible that digital interventions may better support the needs of low-literacy patients through the use of accessible language, opportunities for self-pacing and contextual tips.(77, 78) We were not able to include non-English speaking patients given scope and budget limitations, but if this approach is successful, adaptations for non-English speaking populations is feasible, critically important and a goal for future research.

### Potential for impact and implications

The eREACH2 study is expected to provide critical empiric data on the effectiveness of a theoretically and stakeholder informed digital alternative delivery model in a diverse population of patients who are candidates for germline cancer genetic testing. Equally important, our theoretically-informed process evaluation data is expected to inform modifications to enhance dissemination of our delivery intervention to clinical practice to realize the promise of precision medicine.

## Abbreviations

eREACH: A Randomized Study of an eHealth Delivery Alternative for Cancer Genetic Testing for Hereditary Predisposition in Metastatic Breast, Ovarian, Prostate, and Pancreatic Cancer Patients
GC: genetic counselor
HIPAA: Health Insurance Portability and Accountability Act
MAGENTA: Making Genetic Testing Accessible
PARP: poly (adenosine diphosphate ribose) polymerase
REDCap: Research Electronic Data Capture
SPIRIT: Standard Protocol Items: Recommendations for Interventional
Trials TARGET: Technology-Enhanced Acceleration of Germline Evaluation for Therapy

## Declarations

- **Ethics approval and consent to participate**: The study has been approved by the University of Pennsylvania Institutional Review Board (Protocol# 850242).
- **Consent for publication:** Not applicable
- **Availability of data and materials:** Not applicable
- **Competing interests:** AB has received research funding from AstraZeneca.
- **Funding:** National Cancer Institute U01CA243702 (no role in design, conduct, analysis, or reporting of trial)
- Authors’ contributions
- EM - contributed to visualization and writing—original draft.
- BE contributed to formal analysis, writing—original draft
- KTL contributed to visualization and writing—original draft.
- DM contributed to writing—original draft
- BE contributed to formal analysis, writing—original draft, and writing—review and editing.
- SB contributed to investigation and writing—review and editing.
- SMD contributed to investigation and writing—review and editing.
- KYW contributed to writing—review and editing.
- LW contributed to writing—review and editing.
- JSR contributed to writing—review and editing.
- CC contributed to investigation and writing—review and editing
- JC contributed to investigation and writing—review and editing
- S Howe contributed to investigation and writing—review and editing.
- EMW contributed to investigation and writing—review and editing.
- MW contributed to investigation and writing—review and editing
- KK contributed to project administration and writing—review and editing.
- ES contributed to investigation and writing—review and editing.
- JF contributed to investigation and writing—review and editing.
- SJ contributed to writing—review and editing.
- KS contributed to writing—review and editing.
- BM contributed to investigation and writing—review and editing.
- AB contributed to conceptualization, funding acquisition, investigation, supervision, and writing—original draft.

## Data Availability

No data generated related to this manuscript reporting the study protocol.

## Acknowledgements

KTL’s time was supported by the National Cancer Institute of the National Institutes of Health under award K08CA279076. Generative artificial intelligence was not used in any portion of manuscript generation.

