## Supplemental Tables for "A Randomized Hybrid Type I Effectiveness-Implementation Study of an eHealth Delivery Alternative for Cancer Genetic Testing for Hereditary Cancer (eREACH2): study protocol"

**Supplementary Table 1:** GeM-CheckD: Genomic Medicine Checklist for reporting on Digital tool development, implementation, and evaluation

| **Domain 1: Digital Tool Development** | | | |
|---|---|---|---|
| **Item No.** | **Item Name** | **Description/ sub-Items** | **eREACH** |
| 1.1 | Purpose and Function of the tool | Intent of tool/rationale (e.g., to aid or replace genetic counseling) and function it performs (e.g., help with pre-test or post-test decisions) | To replace traditional pre-test genetic education (visit 1) and/or return of genetic test results (visit 2) with a genetic counselor. Patients still have the option to speak with a counselor. |
| 1.2 | Messaging and Content | Key messages and topics covered by the tool | Visit 1 includes 8 modules which cover purpose of genetic testing and testing options, possible results and implications of results, and testing decision.<br>Visit 2 includes 4 modules which include test results, explanation of result and next steps. |
| 1.3 | Guidelines and/or Theories utilized by the tool | Use of guidelines (e.g., ASCO/ ACMG), frameworks, or models to select content or inform purpose of the tool | Informed by the Tiered-binned model for genetic counseling (Bradbury et al. Gen Med, 2015). |
| 1.4 | Goals/Target of the tool | What is the tool trying to change (e.g., increasing knowledge, reducing decisional conflict, changing behavior) | Provide a digital alternative to reduce the number of scheduled visits for patients and reduce provider time given increasing indications for genetic testing and a limited workforce, while providing equal or better patient outcomes. |
| 1.5 | Intended Users of the tool | Who is the tool for (e.g. patients, providers/ clinicians, general public). May include multiple user types | Patients with cancer where germline testing could inform targeted therapy (breast, ovary, pancreas and prostate cancer). |
| 1.6 | Expertise Leveraged | What experts developed and reviewed the Tool | Genetic counselors, medical oncologists, medical ethicist, behavioral scientists, health communication and medical informatics. |
| 1.7 | Tool Characteristics | Name of the tool. Include version used in project/reporting and the latest version available (if applicable) | eREACH2 Digital Intervention |
| | | Delivery mode (e.g. chatbot, website) | Website |
| | | How information is presented in the tool (e.g. audio/visual, what languages) | Information presented as text, figures, and tables. Optional information includes short videos.<br>Available in English only |
| | | Tool functionalities (e.g. skip logic, interactive, scripted) | Core content is required to be completed by end user. Optional information can be selected to further explore each of the core topics. |
| | | Time for user to complete the tool | ~11-14 mins for visit 1 core (required) content. ~5 mins for visit 2 core content. |
| | | Readability (for written information) | Readability score of $8^{th}$ grade or lower when possible |

| | | | |
|---|---|---|---|
| | | Tailoring/Targeting of content (e.g. adaptation within the tool for different individuals, risks, etc) | Content is customized based on cancer stage (metastatic vs not) and whether there is a known familial mutation. |
| | | Use of AI/LLM within the tool – (specify the type of AI/LLM, closed/open/restricted source, algorithm-based, NLP, etc) | None |
| 1.8 | Development and Refinement Process | Involvement of intended users in developing or refining tool (development and post-development) | Tool refined through user testing with 13 patients and usability testing with 11 patients with cancer. |
| | | Methods used to gather feedback (e.g. interviews, user testing) and any measures used for development and refinement | Individual qualitative interviews for user and usability testing. |
| | | Changes/revisions/ adaptations during development and/or during implementation in context | Changes included: simplifying content, changing order and priority of content, increasing representativeness of photos. |
| **Domain 2: Tool Implementation - Processes and Strategies in Context** | | | |
| **Item No.** | **Item** | **Description/ sub-Items** | |
| 2.1 | Implementation Context | How/when/where tool is accessed by users | Digital intervention is accessed via computer, mobile phone or tablet with a unique user name and password. |
| | | Characteristics of site(s) where tool is deployed | Accessed at home as an alternative to a scheduled visit with a genetic counselor. |
| | | Ethical/governance/considerations for using the tool by a clinic or health system (privacy/safety/ security) | Individual results are included in the disclosure tool requiring user name, password and confirmation of the user. |
| 2.2 | Facilitation | Materials (e.g., manuals, SOPs, support tools) | None. |
| | | Ways of selecting or raising awareness among potential users | Not applicable. |
| | | Instructions on tool use (how, when, where, and how often provided) | Instructions on how to use the tool are included at the beginning of the core content. |
| | | Who facilitates use and how | No facilitation required to complete the intervention. |
| | | Engagement and implementation strategies for end users and facilitators (e.g., prompts, incentives, audit/feedback strategies) | Up to 5 reminders sent to prompt use of the tool |
| 2.3 | Implementation or Process Evaluation and Reporting | Methods for evaluating potential impact of context and/or facilitation on outcomes | CFIR-informed constructs evaluated |
| | | Dose (how much exposure or number of times viewed) and fidelity. Quality Control (QC) process or evaluation to determine if tool is used as intended | Designed to require viewing of at least the Tier 1 information. Analysis of differences in outcomes by use are ongoing. |
| | | Costs involved in implementing the Tool | Not assessed at this time |

| | | | |
|---|---|---|---|
| | | Opinions or experiences of end users or facilitators during implementation | Yes: Study participants, local providers, genetic counselors, and insurers interviewed to inform implementation facilitators, barriers, and adaptations. |
| 2.4 | Implementation implications/ information for scale up or broader use of the tool | Evidence suggesting outcomes may be impacted by context or facilitation | Moderators such as source of care will be collected and evaluated for impact on outcomes. |
| | | Evidence tool may work differently for different groups of people | Moderators of outcomes such as race/ethnicity, gender, and age will be collected and evaluated for impact on outcomes. |
| | | Policy/ practice implications or uses of the Tool broadly | To be determined. |
| | | Potential scalability and maintenance (e.g. prerequisite knowledge and skills to use the Tool, funding source to implement the tool, automatic/manual updating of tool and/or guidelines used in tool) | The tool is mobile. |
| | | Flexibility in implementation (e.g., how easily can the tool adapt to context; are there core elements/processes that can/cannot be adapted) | The tool is not intended to be adapted but changes can be made if needed as clinical care changes. |
| | | Tasks needed for broad dissemination and maintenance | The tool was originally built for only for the research study, but a clinical version is now available upon request. It is also being rebuilt for broader dissemination. |
| | | Remaining gaps in understanding | How time and costs for virtual or digital care will be covered with clinical implementation. |
| **Domain 3: Digital Tool Outcomes Evaluation** | | | |
| **Item No.** | **Item** | **Description/ sub-Items** | |
| 3.1 | Study/ Evaluation Design & Methods | Details about how the effectiveness/ efficacy of the Tool was tested/determined | Randomized non-inferiority trial as described in this manuscript. |
| | | Regulatory/ IRB: how was the implementation and evaluation approved by the IRB to conduct the study and evaluation | Evaluation study was IRB approved. Informed consent was collected upon invitation to use the tool and provide data for evaluation activities. |
| 3.2 | Study/ Evaluation Population | How participants were selected to use the tool (e.g. eligibility criteria) | Patients with breast, ovary, pancreatic or prostate cancer as outlined in this manuscript. |
| | | Incentives provided to participate in the study, complete assessments, or use the tool | No incentive for using the tool. |
| | | Sample size, attrition, participant characteristics | As outlined in the manuscript. |
| 3.3 | Study/ Evaluation Measures & Analyses | Description of outcomes (when measured, what was measured, and what was the primary outcome of the study) | Primary outcomes uptake of genetic testing, knowledge and anxiety. Multiple secondary cognitive, affective and behavioral outcomes and uptake of pre-test |

| | | | |
|---|---|---|---|
| | | | counseling/education and testing. Measured at baseline, after visit 1, after visit 2, and at 6 and 12 months. |
| | | Data handling (e.g. missing data protocols, adjustments, consolidation, transformation, etc) | Not applicable |
| | | Types of analyses conducted | Non-inferiority and moderator analyses as described in this manuscript. |
| 3.4 | Results & conclusions of study/ evaluation using the tool | What do results suggest about the tool's ability to achieve its purpose or goals | To be determined |

**Abbreviations**: ASCO=American Society of Clinical Oncology; ACMG=American College of Medical Genetics; AI=Artificial Intelligence; LLM = Large Language Model; NLP=Natural Language Processing; SOP=Standard Operating Procedure; IRB=Institutional Review Board

**Supplementary Table 2:** Summary of key changes made during development and design of the digital intervention based on user comments during user and usability testing.

| Key changes recommended | Representative user comments |
|---|---|
| User testing | |
| -simplify content | -a lot of information, maybe use more bullets (V1)<br>-Keep it simple, especially if someone already has it--getting to technical here- could lose a lot of people--want more focus how it can help you (V1)<br>-might be a little too much, possibly split into 2 slides (V2) |
| -address confusing content | -word somatic confuses initially, nice to be able to get defintions of words when you hover over (easier and faster than glossary)(V1)<br>-switch between inherited and germline confusing-make the order consistent (V1)<br>-what family members? Children, siblings, parents or distant? (V2) |
| -address benefits of testing equally to risks and limitations | -Too much info on uncertainty. Will just make people nervous (V1) |
| -add more information on costs and consider addressing earlier | -This info needs to be clear earlier. Could be a big barrier. Maybe something about speaking to a gc with concerns (V1)<br>-0-100 I can afford, should be higher up in the presentation, it is important to know (V1) |
| -move content to Tier 2 or Tier 1 | -[example case scenario] doesn't mean anything to me at this point- not useful or helpful- should be extra category (V1)<br>-needs to be in the extra category- might make people's eyes glaze over (V1) |

| | |
|---|---|
| | -reinforces decision making – thinks this should be in tier 1 (V1) |
| Usability testing | |
| -clearer instructions and labeling | -unclear how to advance through a set of 4 screens – make arrows more clear (V1)<br>-have a back arrow in case someone changes their mind and doesn't want to proceed to results (V2)<br>-placement of the help button is confusing (V2) |
| -functionality | -videos are slow (V1)<br>-page did not load (V2) |
| -changes in size of text and figures | -diagram is too small to read (V1)<br>-text is too small or should be bold (V2) |
| -changes to images | -photo of physician is intimidating (V1)<br>-need more photos of older people (V2) |
